# A novel syndrome caused by the constitutional gain-of-function variant p.Glu1099Lys in *NSD2*

**DOI:** 10.1101/2022.02.23.22271353

**Authors:** Bernt Popp, Melanie Brugger, Sibylle Poschmann, Tobias Bartolomaeus, Maximilian Radtke, Julia Hentschel, Nataliya Di Donato, Andreas Rump, Janina Gburek-Augustat, Elisabeth Graf, Matias Wagner, Johannes Lemke, Thomas Meitinger, Rami Abou Jamra, Vincent Strehlow, Theresa Brunet

**Affiliations:** Institute of Human Genetics, University of Leipzig Medical Center, Leipzig, Germany; Institute of Human Genetics, Klinikum rechts der Isar, School of Medicine, Technical University of Munich, Munich, Germany; Division of Neuropediatrics, Clinic for Children and Adolescents Dritter Orden, Munich, Germany; Institute for Clinical Genetics, University Hospital, TU Dresden, Dresden, Germany; Division of Neuropaediatrics, Hospital for Children and Adolescents, University of Leipzig Medical Center, Leipzig, Germany; Institute of Neurogenomics, Helmholtz Zentrum München, Neuherberg, Germany; Division of Pediatric Neurology, Developmental Medicine and Social Pediatrics, Department of Pediatrics, Dr. von Hauner Children’s Hospital, Munich University Hospital (Ludwig Maximilians University), Munich, Germany

**Keywords:** NSD2, gain-of-function, Glu1099Lys, neurodevelopmental disorder, Rauch-Steindl syndrome, Wolf-Hirschhorn syndrome

## Abstract

**Purpose:** NSD2 dimethylates histone H3 at lysine 36 (H3K36me2) and is located in the Wolf-Hirschhorn syndrome (WHS) region. Recent descriptions delineated loss-of-function (LoF) variants in *NSD2* with a distinct disorder. The oncogenic missense variant p.Glu1099Lys occures somatically in leukemia and has a gain-of-function (GoF) effect.

**Methods:** We describe two unrelated individuals carrying c.3295G>A, p.Glu1099Lys as heterozygous *de novo* germline variants identified by exome sequencing of blood DNA and subsequently confirmed in two ectodermal tissues. We use omics data from the Cancer Cell Line Encyclopedia to analyze the GoF effect.

**Results:** Clinically these individuals are characterized by intellectual disability, coarse/ square facial gestalt, abnormalities of the hands and organomegaly. We confirmed increased K36me2 methylation in lines with either *NSD2* GoF variants or duplications. Cells with GoF variants showed increased DNA promoter methylation and dysregulated RNA expression, influencing cellular modules involved in white blood cell activation, cell growth and organ development.

**Conclusion:** NSD2 GoF caused by the missense variant p.Glu1099Lys leads to a novel syndromic phenotype distinct from both the previously described LoF phenotypes. Other variants causing NSD2 hyperactivation or overexpression may cause a similar phenotype. This syndrome should be distinguished from the recently named Rauch-Steindl syndrome caused by *NSD2* haploinsufficiency.

## INTRODUCTION

Variants affecting genes encoding proteins involved in epigenetic processes are a prominent cause for neurodevelopmental disorders (NDDs) and have recently been summarized as “Mendelian Disorders of the Epigenetic Machinery”.^1^ Clinically, these entities often comprise intellectual disability (ID) and growth abnormalities.

Post-translational modifications (acetylation or methylation) of lysine (K) residues in the N-termini of histones are among the most common mechanisms of epigenetic regulation in the cell. These modifications can determine the accessibility of DNA and therefore its transcriptional activity. They mainly affect the H3 core histone and, depending on the modified amino acid (AA) residues, can serve as a marker for repressive (like H3K27) or active (like H3K36) chromatin states.

The nuclear receptor binding SET domain protein 2 (NSD2) catalyzes writing of H3K36 methylation marks.^2,3^ The previous name “Wolf-Hirschhorn syndrome candidate 1” (*WHSC1*) indicates that the gene is located in the Wolf-Hirschhorn syndrome (WHS, OMIM #194190) deletion region on the short arm of chromosome 4. WHS has been cytogenetically characterized prior to the chromosomal microarray (CMA) era and subsequently refined as a contiguous gene deletion syndrome with two candidate genes (*NSD2* and *LETM1*) in a 1.6 Megabase (Mb) critical genomic region on 4p16.3 (OMIM #194190). The core clinical features of WHS include besides NDD a typical facial dysmorphology described to resemble a “Greek warrior helmet”, ID and intrauterine/ postnatal growth retardation.^4,5^ With the broad availability of exome sequencing, individuals with NDDs carrying heterozygous *NSD2* variants have been reported subsequently.^6–8^ A recent article described truncating and missense variants in *NSD2* in 18 individuals from 16 families with a rather mild developmental phenotype that only partially overlaps with WHS (Rauch-Steindl syndrome, RAUST, OMIM #619695).^9^ Functional analyses of the five different missense variants showed decreased ability to generate H3K36me2 in all but one (c.2606G>A, p.(Cys869Tyr)), indicating that loss of methylation activity is the main pathomechanism.^9^

The other previous name of *NSD2*, “multiple myeloma SET domain containing protein” (MMSET), highlights the gene’s involvement in hematologic malignancies, which was initially identified by characterization of the somatic t(4;14) translocation present in 15-25% of multiple myeloma cases.^10^ This rearrangement fuses *NSD2* to the immunoglobulin heavy-chain (IGH) and thus causes its overexpression. Sequencing studies later identified a recurring somatic hotspot mutation in *NSD2* (E1099K; c.3295G>A, p.Glu1099Lys) in cases of pediatric acute lymphoblastic leukemia.^11,12^ Functional studies showed that the IGH-NSD2 fusion causes expansion of H3K36me2 methylation marks and differential gene expression with oncogenic potential.^13^ Similarly the p.Glu1099Lys oncogenic missense variant within *NSD2* enhanced global H3K36me2 in lymphoid cells and reduced H3K36 trimethylation.^11^ Recent investigations using cryo-electron microscopy solved the structure of NSD2 bound to the nucleosome and indicated that the oncogenic p.Glu1099Lys *NSD2* missense variant destabilizes the autoinhibitory loop of the protein to enhance substrate recognition and increase the nucleosomal H3K36 methyltransferase activity of NSD2.^14,15^

It is known ^16^ that genes involved in clonal hematopoiesis, hematologic malignancy and other types of cancer are also overrepresented in curated NDD databases. On a variant level, there is a significant overlap between somatic driver mutations in (hematologic) malignancies and (*de novo*) germline variants causing NDD. Both *de novo* missense variants identified in large NDD cohorts and somatic driver variants in cancer ^17^ are enriched in constrained genomic regions. These observations seem plausible, because many of the overlapping genes are central to cellular processes and similar mutational mechanisms affect both somatic and germline cells.

Prominent examples for cancer associated genes with two distinct associated NDD entities based on different molecular alterations caused by the underlying variants include epigenetic regulators like *DNMT3A* (OMIM #618724, “Heyn-Sproul-Jackson syndrome”; OMIM #615879, “Tatton-Brown-Rahman syndrome” ^18^) and *SETD2* (OMIM #616831, “Luscan-Lumish syndrome”; a clinically distinguishable disorder associated with GoF missense variants at AA position Arg1740 recently described by Rabin and colleagues ^19^). Likely further similar entities caused by distinct functional effects of disease-causing variants in the same gene are to be discovered.

Here, we describe the identification of the GoF variant p.Glu1099Lys in *NSD2* as a constitutional, germline heterozygous *de novo* variant in two individuals with a clinically recognizable syndromic NDD entity.

## MATERIALS AND METHODS

### Ethics approval and consent to participate

The Ethical Committee of the Medical Faculty, Leipzig University, approved genetic testing in a research setting for individual Ind_1 (224/16-ek and 402/16-ek). For individual Ind_2, the study was approved by the local ethics committee of the Technical University of Munich (#5360/12S). Written consent of the parents of both individuals to publish genetic, clinical data and images was received and archived by the authors.

### Genetic analyses and comparison of phenotypes and genotypes

Variants in the index cases were identified by trio exome sequencing (ES) as described previously ^20,21^. We used an Excel based questionnaire for retrospective phenotyping of the individuals described here and for review of published cases. Details regarding ES, including primer sequences for Sanger sequencing using DNA from three different tissues, and the clinical sheet are provided in Supplementary notes.

### Variant spectrum and 3D structure analysis

The *NSD2* germline variants were harmonized to a common reference with VariantValidator ^22^ (NM_001042424.2 transcript, hg19 reference). The distribution of NDD associated *NDS2* variants in the linear protein representation and in the tertiary protein structure (PDB 7CRO ^14^) was compared to variants from databases as described previously ^23^ (details in Supplementary notes).

### Analysis of omics data from cancer cell lines

We analyzed data from the Cancer Cell Line Encyclopedia (CCLE) ^12^ using “depmap ^24^ the ExperimentHub R-package regarding mutation and copy number calls, chromatin modifications, differential methylation and differential expression. Humanbase ^25^ (https://hb.flatironinstitute.org/) was used to predict functionally enriched gene modules. Compare File S2 and Supplementary notes for details.

## RESULTS

### Genetic analyses and review of *NSD2* variants

Trio exome sequencing detected a heterozygous *de novo* missense variant in *NSD2* (NM_001042424.3: c.3295G>A, p.Glu1099Lys; chr4[hg19]:g.1962801G>A) in both individuals described here. Constitutional status of the c.3295G>A, p.Glu1099Lys *NSD2* variant was confirmed through Sanger sequencing of DNA from buccal swabs and fingernails in Ind_1 or from buccal swabs in Ind_2 (Fig. S01).

The *NSD2* variant c.3295G>A, p.Glu1099Lys, had not been previously described as disease-causing in the literature when mutated in the germline and was listed once (1/251,470 alleles) in gnomAD in an individual assigned to the age group of 75-80 years. However, when visualizing the variant listed in gnomAD, it was present in only 32/89 reads (variant allele fraction of 36%) indicating somatic mosaicism (compare Fig. S02). As the phenotype of the individuals did not match with LoF descriptions (RAUST or WHS) the variant was classified as uncertain significance (VUS) according to the ACMG (American College of Medical Genetics and Genomics) recommendations ^26^ applying the criteria (PS2_moderate, PP3, PM2_supporting). Based on the significant clinical overlap of the two individuals, we upgraded the ACMG criterion PS2_moderate to PS2_strong and added PM1_strong (multiple functional analyses) and PP2 (missense Z-score=3.9), allowing classification as “pathogenic”.

Previously described LoF variants affecting *NSD2* are dispersed throughout the protein (Fig. 2A). The distribution in samples from the COSMIC database is similar and shows peaks for the recurrent GoF variants (Fig. 2A). Missense variants with a presumed LoF effect are located in functional domains, but show no clustering in the tertiary protein structure while the two missense variants with a proven GoF effect are relatively close to each other and the lysine at AA position 36 of H3 (Fig. 2B).

**Figure 1.**
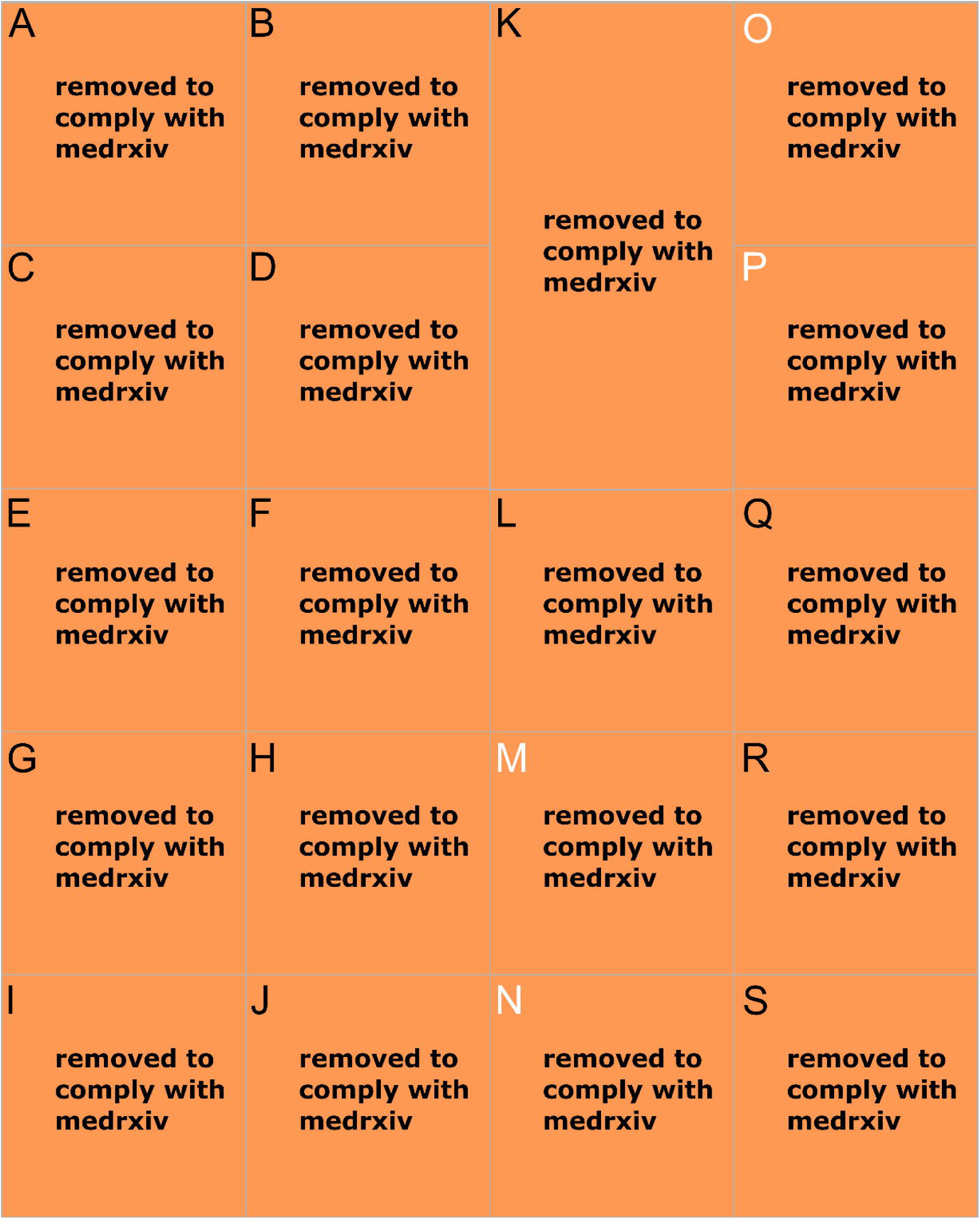
Facial dysmorphology and clinical images. **(A)** and **(B)** frontal facial images of Ind_1. **(C)** and **(D)** lateral facial images of Ind_1. facial images of Ind_1. **(E)** and **(F)** dorsal and **(G)** and **(H)** palmar sides of the right hand of Ind_1. **(I)** and **(J)** dorsal side of the feed of Ind_1. Paired images at different ages. **(K)** Full frontal body view of Ind_1. **(L)** Hip X-ray of Ind_2 showing almost horizontal acetabulum, widened proximal femoral metaphysis and prominent incisura ischiadica indicative of skeletal dysplasia. **(M)** and **(N)** abdominal MRI images of Ind_2 showing hepatomegaly and nephromegaly. **(O)** abdominal ultrasound of Ind_2 displaying kidney hypertrophy, diminished corticomedullary differentiation, irregular parenchyma, multiple small renal cysts. and **(O)** hepatomegaly with rounded caudal margin in Ind_2. **(Q)** and **(R)** Dorsal and palmar sides of the right hand of Ind_2 showing short and tapered fingers as well as hypoplastic fingernails. **(S)** dorsal side of the feet of Ind_2 illustrating hypoplastic toenails.

**Figure 2.**
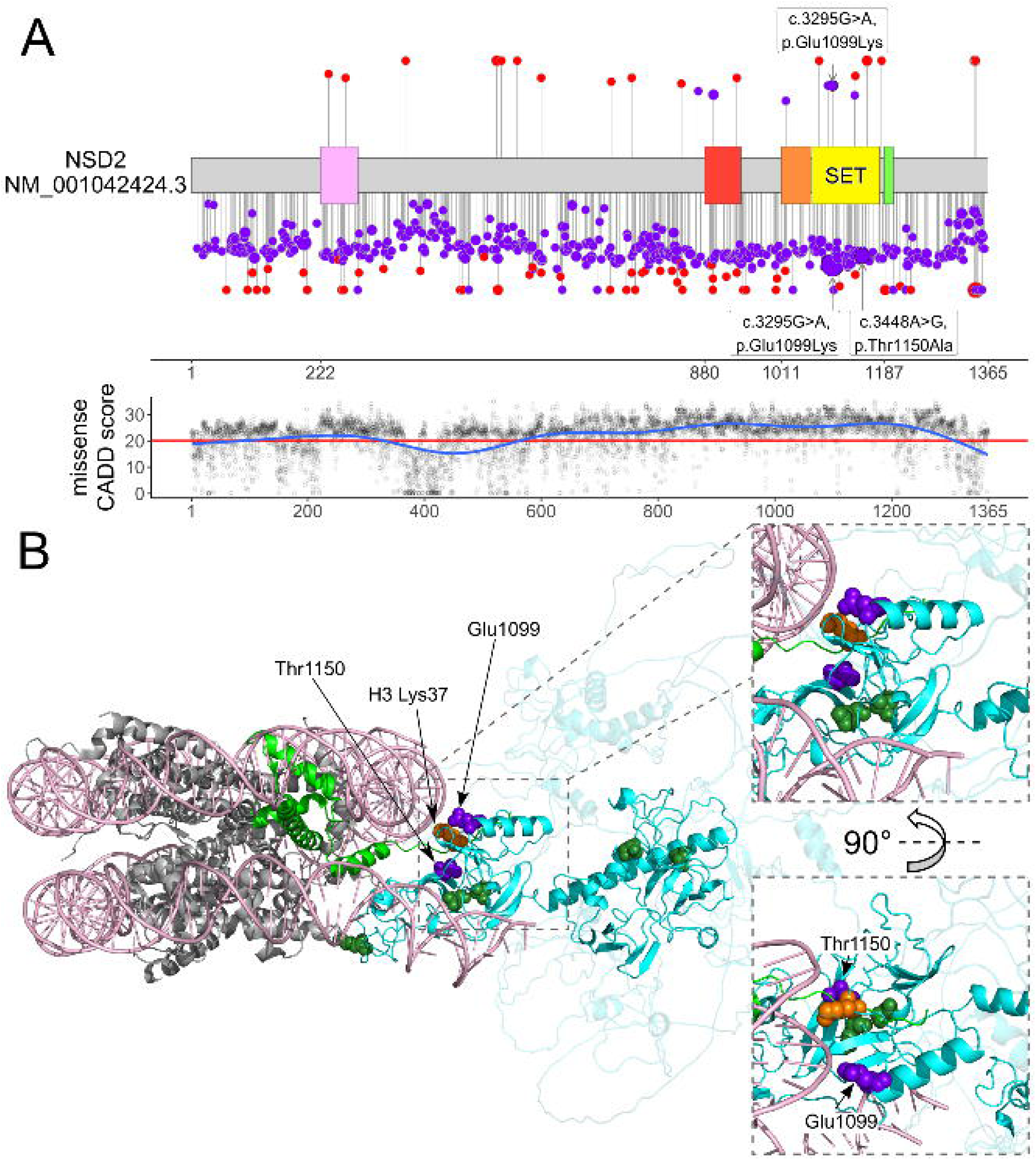
Variant distribution on NSD2. **(A)** Upper panel with schematic of the linear NSD2 protein with domains and variant distribution (missense variants in blue, truncating variants in red). Variants above the protein scheme have been described in germline, while the ones below are somatic from the COSMIC database (note the GoF variant occurring 50 times). The position of the p.Glu1099Lys GoF variant reported here is indicated by the label and arrow. The size of the lollipop circle is proportional to the reported frequency in the literature or database. The segment height is proportional to the CADD-score. Domains based on UniProt O96028. Lower panel with the generalized additive model of the CADD PHRED v1.6 values for all possible NSD2 missense variants as indicator of conserved protein domains indicates highest constraint in the SET domain. **(B)** Pseudo model of the nucleosome with DNA double helix (light pink), histone octamer (gray) and NSD2 (cyan) bound to H3 (green). The less structured regions of NSD2 (amino acid (AA) positions 1-760 and 1200-1365) are faded. The position of AA affected by missense variants in NSD2 from our study (Glu1099) are presented as blue and from the literature review as green spheres. The AA of the second described GoF missense (Thr1150) variant is also colored in blue. The H3 lysine 36 (K36) is colored in orange. Right panels show zooms of the K36 region (upper right side) and a view from below the model rotated by 90° (lower right side). Overall, the likely LoF missense variants are further away from K36 and more dispersed throughout the NSD2 protein.

### Comparing NSD2 allelic disorders

Both individuals described here showed a remarkable phenotypic overlap with complicated pregnancies and postnatal periods, prenatally detectable organomegaly and postnatally evident dysmorphic features characterized by coarse and rather square facial features (Fig. 1B-D, K) and large hands with tapering fingers (please compare the Supplementary notes for detailed clinical case reports of Ind_1 and Ind_2). Noticing this phenotypic overlap we wanted to attempt a genotype-phenotype correlation in comparison with published LoF cases.

The literature review identified 26 individuals with (likely) pathogenic and presumed LoF variants from six publications in which case specific phenotype data was reported.^6–9,27,28^ They carried 23 different variants, mostly truncating (n=18, 78.3%) and few missense (n=5, 21.7%). As the descriptions were reported in different publications we first normalized and reviewed the clinical descriptions to compare these with the two cases identified in our study carrying the GoF variant c.3295G>A, p.Glu1099Lys. Despite the sample size of two for the GoF group and the LoF group being inhomogeneous, we identified several facial dysmorphologies significantly varying. These include the coarse facial features with a rather square face in the GoF cases compared to the rather triangular face of individuals with LoF variants. The LoF cases had a broad forehead with high anterior hairline, while the low anterior hairline reported on both GoF cases indicated a smaller forehead area. Facial features exclusively reported in both the GoF cases included anteverted nares and an exaggerated cupid’s bow, while a long philtrum was described on both GoF and 1/15 LoF cases. While 6/20 facial features reached nominal significance, due to the small sample size no features remained significant after correction for multiple testing (Table 1).

**Table 1.** Phenotypic abnormalities of the face.

| Phenotype | HPO | GoF cases | LoF cases | p-value (FDR-corr.) |
| --- | --- | --- | --- | --- |
| Abnormal facial shape | HP:0001999 | 100% (2/2) | 100% (16/16) |  |
| **Coarse facial features** | **HP:0000280** | **100% (2/2)** | **12% (2/16)** | **0.039 (0.189)** |
| Triangular face | HP:0000325 | 0% (0/2) | 81% (13/16) |  |
| **Square face** | **HP:0000321** | **100% (2/2)** | **0% (0/16)** | **0.007 (0.090)** |
| Thick hair | HP:0100874 | 100% (2/2) | 0% (0/1) |  |
| High anterior hairline | HP:0009890 | 0% (0/2) | 80% (12/15) |  |
| **Low anterior hairline** | **HP:0000294** | **100% (2/2)** | **13% (2/15)** | **0.044 (0.190)** |
| Broad forehead | HP:0000337 | 0% (0/2) | 81% (13/16) |  |
| Laterally sparse eyebrows | HP:0005338 | 50% (1/2) | 53% (8/15) |  |
| Large palpebral fissures | HP:0001090 | 50% (1/2) | 67% (10/15) |  |
| Hypertelorism | HP:0000316 | 50% (1/2) | 37% (7/19) |  |
| Periorbital hyperpigmentation | HP:0001106 | 0% (0/2) | 86% (12/14) |  |
| Infra-orbital crease | HP:0100876 | 100% (2/2) | 33% (4/12) |  |
| **Anteverted nares** | **HP:0000463** | **100% (2/2)** | **0% (0/15)** | **0.007 (0.090)** |
| Wide nasal bridge | HP:0000431 | 100% (2/2) | 39% (7/18) |  |
| **Long philtrum** | **HP:0000343** | **100% (2/2)** | **7% (1/15)** | **0.022 (0.136)** |
| Short philtrum | HP:0000322 | 0% (0/2) | 40% (6/15) |  |
| Thin upper lip | HP:0000219 | 100% (2/2) | 35% (6/17) |  |
| **Exaggerated cupid's bow** | **HP:0002263** | **100% (2/2)** | **0% (0/14)** | **0.008 (0.090)** |
| Posteriorly rotated ears | HP:0000358 | 50% (1/2) | 80% (4/5) |  |

When comparing other phenotype categories, a short neck was present in both GoF cases but in 0/12 LoF cases with data. Tapered fingers were described in both GoF cases but in only 1/18 LoF cases. Remarkably, four other hand phenotypes were described in both GoF cases, but were not phenotyped in the LoF cases (Table 2; compare Fig. 1E-H and Q-R).

**Table 2.**
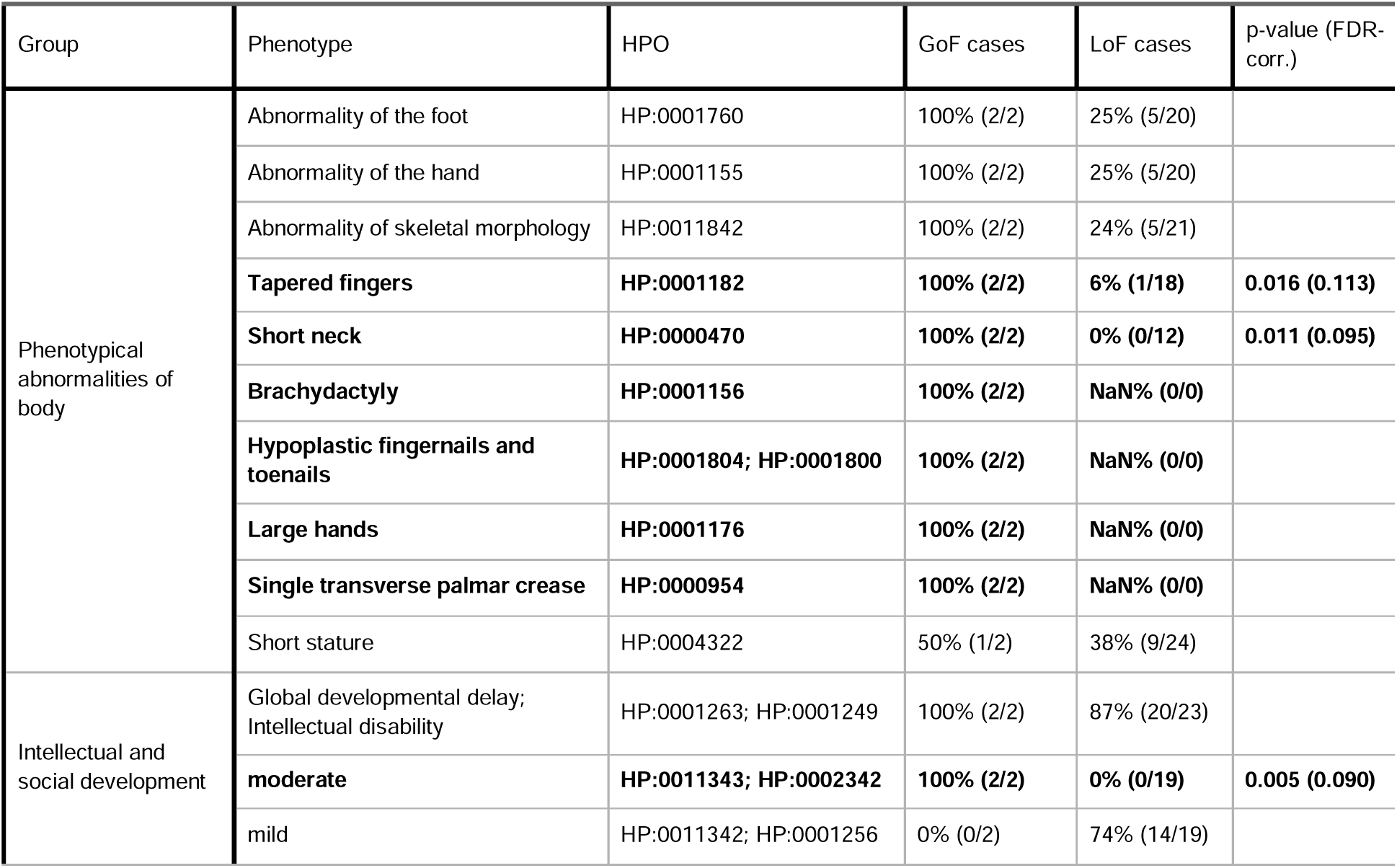
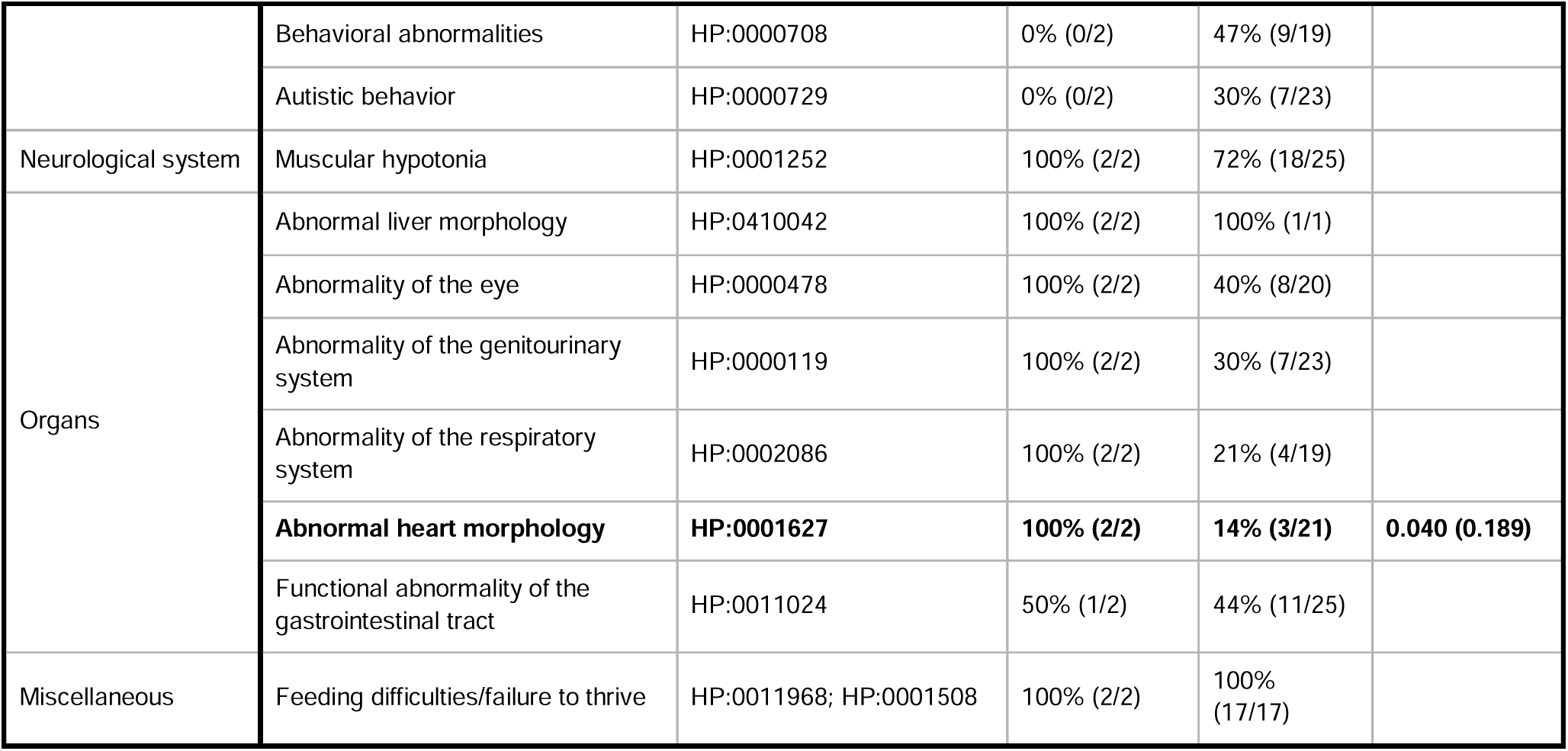
Comparison of other phenotypes between of GoF and LoF cases.

Regarding organic abnormalities only abnormal heart morphology reached nominal significance, but the patent ductus arteriosus in Ind_1 is a relatively nonspecific and frequent finding which indicates that the associated phenotype needs to be refined in further descriptions. Abnormalities of the genitourinary system were described in both GoF cases but also in 7/23 (30.4%) of LoF cases. It should be noted that abnormal liver morphology was present in both GoF cases but data on this phenotype was only reported for one LoF; assuming that the other LoF cases were healthy in this regard, this feature would also be nominally significant. Intellectual disability or developmental delay was reported as moderate in both GoF cases, while most LoF cases had mild intellectual disability (73.7%; 14/19). Behavioral anomalies (47.4%; 9/19) and autistic behavior (30.4%; 7/23) were reported in the LoF cases but absent in the two GoF cases. Again, 4/23 phenotypic features analyzed reached nominal significance but did not withstand correction for multiple testing (Table 2).

### Altered H3K36 modification and DNA methylation changes in NSD2 GoF cell lines

Our CCLE ^12^ analysis of global chromatin modification showed that cell lines with GoF variants in *NSD2* had a significantly higher mono-(me1) and dimethylation (me2) level at H3 lysine 36 (K36) with mirroring significant reduction of unmethylated (me0) K36 (Fig. 3A). Cell lines with a genomic duplication affecting the *NSD2* gene region, similarly showed increased ratios of K36me1 and K36me2 with reduction of K36me0, with a lesser effect (Fig. 3B). Differential DNA methylation analysis leveraging the CCLE omics dataset identified a large fraction of promoter regions being differentially methylated. Most loci had a small depmap effect size between the GoF and *NSD2* variant neutral groups and most loci were hypermethylated (n=5,165) while only few showed DNA hypomethylation (n=117). When comparing large depmap effect sizes (defined as > 0.5 or < -0.5) only 12 loci were hypermethylated compared to 28 hypomethylated (Fig. 3C). Of the promoter loci showing a high hypermethylation effect, the *TTC12* (“Ciliary dyskinesia, primary, 45”, OMIM #618801) and the *NR2F2* (“46,XX sex reversal 5”, OMIM #618901; “Congenital heart defects, multiple types, 4”, OMIM #615779) are associated with human disease in the OMIM database. The *NR2F2*-disorders are associated with congenital heart defects and abnormalities of genitalia. The genes *KPNA7* and *CDC42BPB* have the two most highly hypermethylated promoters when considering NDD-associations curated in the SysID ^29^ database (compare File S2 sheet “GoFvsNeutral_Methylation”).

**Figure 3.**
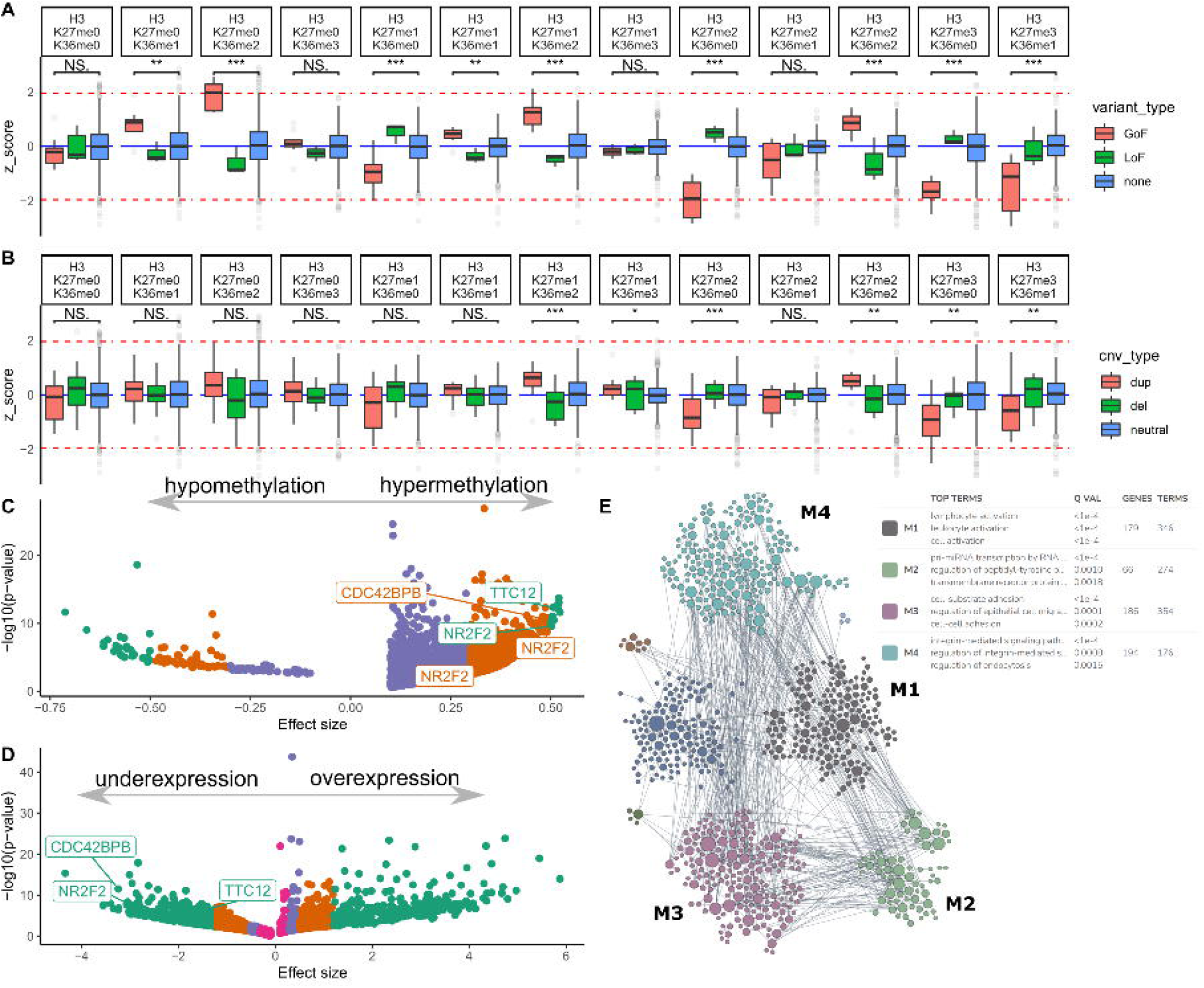
NDS2 and cell line omics analyses. **(A)** Boxplots comparing the z-scores of the global chromatin profiling dataset (CCLE) between cell lines with GoF (red), LoF (green) and no (blue) *NSD2* variant for modification combinations at histone H3 at K27 and K36. Results show a strong hypermethylation effect with increased mono- and dimethylation at K36 and associated reduction of unmethylated (me0) K36 for cell lines with GoF. Cell lines with a LoF variant showed a trend towards an opposite effect on histone modification but did not reach statistical significance (comparison bars not shown), which could be explained by the weaker effect of copy number changes (compare duplications) or other genetic effects in the control cell lines (e.g promotor or non-coding variants) causing similar changes in histone modification. **(B)** Boxplots as in (A) but now comparing cell lines with genomic duplications (“dup”, red), deletions (“del”, green or copy neutral (blue) state at the NSD2 gene locus. The results for duplications mirror the observations for GoF in (A) but with a weaker effect. (two sided Wilcoxon signed-rank test; NS, not significant; “***”, 0.001; “**”, 0.01, “*”, 0.05). **(C)** Scatterplot of the differential DNA methylation analysis between cell lines with GoF and no NSD2 variant using the reduced representation bisulfite sequencing (RRBS) dataset of methylation one kilobase upstream of the transcription start site (TSS1kb). Only genetic loci with an effect size > 0.1 or < -0.1 are displayed and effect sizes are color coded. Overall, most differentially methylated DNA loci are hypermethylated (5,165 vs. 117 with hypomethylation). When comparing large effect sizes (> 0.5 or < -0.5) only few loci are hypermethylated (12 vs. 28 hypomethylated). **(D)** Scatterplot of the differential RNA expression using the same cell line lists as in (C) shows many genes dysregulated with a very large effect size (887) with 311 showing overexpression and 576 with reduced expression. Dysregulated gene loci with disease association are highlighted by a text box (note that *NR2F2* has different start sites and thus multiple promoter regions marked). Analyses in C and D have been performed using the depmap portal “Data Explorer” tool. **(E)** Network map of functionally enriched modules in the 887 genes with very high effect sizes from (D). The four largest modules (M1: 179 genes, M2: 66 genes, M3: 186 genes, M4: 194 genes) are highlighted with their top associated terms in the accompanying box. The modules are enriched for white blood cell activation (M1), cell adhesion and growth (M2, M3, M4) and organ development (M2, M3).

### Dysregulated pathways in NSD2 GoF cell lines

Differential RNA expression analysis using the sets of NSD2 GoF and neutral cell lines resulted in a large set of dysregulated genes (n=2,284 with q-value < 0.05; Fig. 3D). Considering only genes with a very large depmap effect size (n=887; defined as > 1.2 or < - 1.2), 311 showed overexpression and 576 showed a reduced expression. The OMIM disease genes *TTC12* and *NR2F2* identified to have a hypermethylated promoter region were also in the downregulated gene set with very high depmap effect size. Similarly, *CDC42BPB*, identified in the SysID overlap analysis and recently associated with NDD ^30^, was in the group of genes with very high depmap effect size. Less than expected (n=54; p-value ∼ 0.035; two sided Binomial test) NDD genes from SysID (status November 18th 2021 with 1534 primary genes; 1523 present in the depmap expression dataset) overlapped with the very high effect expression geneset (compare File S2 sheet “GoFvsNeutral_Expression”).

Finally, we sought to functionally cluster the 887 genes with a very high effect on expression to explore dysregulated cellular mechanisms. The humanbase analysis identified eight functionally enriched modules with four of them (M1-4) containing most of the genes (Fig. 3E). The M1 module comprised 179 genes and was significantly enriched for blood cell activation terms (e.g. top two terms: “lymphocyte activation”, GO:0046649; “leukocyte activation”, GO:0045321). The M2 module contained 66 genes enriched for tyrosine kinase pathway (e.g. term number two “regulation of peptidyl-tyrosine phosphorylation”, GO:0050730) and kidney development (e.g. term number four “positive regulation of metanephros development”, GO:0072216). Module M3 contained 186 genes with an enrichment cell adhesion (top term “cell-substrate adhesion”, GO:0031589) and growth terms (e.g. term four “hippo signaling”, GO:0035329). Module M4 contained 194 genes enriched for cellular growth (top term “integrin-mediated signaling pathway”, GO:0007229) and embryonic development (e.g. term four “gastrulation”, GO:0007369). Overall, the dysregulated modules are in line with the known function of NSD2 GoF in leukemia and the organ overgrowth phenotype in the two individuals described here.

## DISCUSSION

A genetic disorder can be described most unambiguously by the affected gene, the proposed inheritance pattern, the phenotypic description of associated disease and finally the precise effect of pathogenic variants on protein function. This differentiation is important to guide standardized genetic diagnostics to enable well-characterized rare disease cohorts. *NSD2* was previously recognized as a candidate gene in the WHS region and truncating/ missense variants with a loss-of-function (LoF) effect in *NSD2* had been associated with a milder NDD entity with distinct facial appearance (Rauch-Steindl syndrome; RAUST). Thus, *NSD2* haploinsufficiency explains only part of the WHS phenotype.

Next to these entities, the *NSD2* missense variant p.Glu1099Lys has been extensively studied, because of its somatic recurrence in leukemia caused by a gain-of-function (GoF) effect. Here, we report two individuals with a syndromic NDD presentation harboring the germline GoF variant c.3295G>A, p.Glu1099Lys in *NSD2*. Comparing the facial gestalt of individuals with WHS is complicated as further genes in the deletion are expected to contribute to some of the most distinguishing features like the wide nose bridge devolving into the forehead (“Greek warrior helmet”), but there are some overlaps with individuals carrying *NSD2* LoF variants such as high anterior hairline, broad forehead, arched eyebrows, large palpebral fissures, short philtrum. Notably, when comparing the described individuals with NSD2 LoF individuals, some facial features seem inverted. While GoF carriers have a square facial appearance with low anterior hairline and small forehead, LoF carriers have a triangular face with high anterior hairline and large appearing forehead. Similarly the nasal bridge is wide and depressed on the two individuals with the c.3295G>A, p.Glu1099Lys variant while a thin and elevated nasal bridge was described as specific in the LoF cohort. With the organomegaly and the abnormalities of the skeleton and hands/ feet, the appearance of the NSD2 GoF individuals seems rather syndromic compared to the relatively unspecific presentation in the LOF individuals.

These phenotypic dissimilarities are expected, given the opposite cellular effects of GoF and LoF variants. Utilizing publically available omics data from the Cancer Cell Line Encyclopedia (CCLE) ^12^ we confirm ^15,31^ dysregulated methylation of histone H3 at the lysine residue 36, especially increased dimethylation, (H3K36me2) as primary molecular effect of the c.3295G>A, p.Glu1099Lys variant. Looking at differential promoter methylation, we identified extensive DNA hypermethylation in GoF cell lines (Fig. 3). The DNA-(cytosine-5)-methyltransferase enzymes (DNMTs) recognize H3K36me2 with their PWWP (AAs ‘Pro-Trp-Trp-Pro’) domain and bind to these regions of open chromatin.^32^ This effect of NSD2 on DNA methylation can be used in future studies to generate specific epi-signatures from peripheral blood DNA, which could aid to characterize variants.

As DNA methylation is involved in gene expression, we next investigated RNA data from the CCLE, which identified >800 genes differentially expressed with very large effect size between GoF and LoF cell lines. Interestingly, less than expected genes currently associated with NDDs in the SysID databse overlapped with these dysregulated geneset. This observation however is in line with the relatively mild developmental phenotype observed in Ind_1 (brain hemorrhage and hypoxia complicated by the course of Ind_2), considering the genome wide effect of the *NSD2* GoF variant c.3295G>A, p.Glu1099Lys. Using the humanbase tool ^33^ to cluster the dysregulated geneset, we identified eight main functional modules with the largest and most significantly enriched four being involved in white blood cell activation, cell growth and organ development (Fig. 3E). These enriched functions overlap well with known cellular functions of NSD2 and with the phenotype observed in carriers of the c.3295G>A, p.Glu1099Lys GoF variant. Other “Mendelian Disorders of the Epigenetic Machinery” entities like Sotos syndrome (OMIM #606681; *NSD1* gene), Tatton-Brown-Rahman syndrome (OMIM #615879; *DNMT3A* gene), and Luscan-Lumish syndrome (OMIM #616831; *SETD2* gene) are also associated with overgrowth symptoms like macrocephaly and tall stature.

While the c.3295G>A, p.Glu1099Lys variant in Ind_1 was identified by clinical ES about six years prior to this report, its highly suspected pathogenicity remained unclear until a second case bearing the same variant was identified. This anecdotal diagnostic odyssey for Ind_1 together with no other germline descriptions of this variant, despite the relatively large *NSD2* cohort ^9^ collected through matchmaking, argues for a very rare disorder. The description of this rare entity was enabled through the practice of openly sharing all diagnostic classifications through ClinVar. Another possible explanation of this association being described after the LoF variants, is that the c.3295G>A, p.Glu1099Lys *NSD2* variant is a known somatic variant in leukemia, which is present in one elderly individual from gnomAD, and could thus be excluded in some very strict filtering pipelines^34^.

Next to the p.Glu1099Lys missense substitution, other *NSD2* missense variants causing similar GoF effects have either been described in blood malignancies (T1150A; c.3448A>G, p.Thr1150Ala) or investigated in vitro ^14,15^. Also structural variants like the recurrent somatic t(4;14) translocation, can result in overexpression and increased activity of NSD2. This is intriguingly confirmed by our observation that genomic duplications in cell lines cause a similar hypermethylation profile to GoF variants (Fig. 3B), which indicates that NSD2 could be triplosensitive and contribute to the not well defined 4p16.3 microduplications.

Due to the rarity of this novel NSD2 GoF entity it is hard to predict the clinical outcome and give recommendations regarding treatment and surveillance. Both individuals with the GoF variant required intensive care postnatally. The course of Ind_2 was complicated by brain hypoxia, which hampers assessing his development. Ind_1 on the other hand shows a mild to moderate intellectual disability, which seems surprising given the strong histone methylation changes induced by the GoF variant. Similarly, while both individuals had organ abnormalities and organomegaly (kidneys, liver), laboratory findings were within normal ranges indicating no signs of beginning organ failure. Due to the strong association of the c.3295G>A, p.Glu1099Lys variant with leukemia, the most obvious concern in the two cases remains a possibly increased blood cancer risk, despite unremarkable repeated blood counts. An increased cancer risk has been described in other “Mendelian Disorders of the Epigenetic Machinery” caused by certain variants in *NSD1, DNMT3A, SETD2* and others.^35– 37^ A murine conditional knock-in model of the p.Glu1099Lys variant in the b-cell lineage did not lead to spontaneous development of leukemia in immunocompetent mice during the expected life span.^31^ Enrichment in relapses of acute lymphoblastic leukemia and functional studies showing induction of glucocorticoid resistance indicate that the p.Glu1099Lys NSD2 GoF variant might cause clonal advantage and not act as a primary driver event.^38^ Based on our current knowledge, we would still recommend at least yearly blood counts, coupled with the regular developmental assessment and abdominal organ ultrasound. Additionally, efforts are made to develop small molecule inhibitors targeting NSD2 as a novel drug in the context of hematologic malignancies ^39,40^ posing a ground to investigate therapeutic strategies in GoF germline carriers.

Here, we describe a syndromic NDD entity associated with increased enzymatic activity of the *NSD2* gene product causing disturbed histone methylation and dysregulation of gene networks correlating with the observed phenotypes of organomegaly. This relatively specific phenotype, coupled with recognizable coarse facial features and abnormalities of the hands, may allow the clinically supported diagnosis of this “Mendelian Disorders of the Epigenetic Machinery” in individuals with other *NSD2* variants identified by ES. Due to the clinical distinguishability from the previously described syndromes associated with genomic loss of the NSD2 gene region (WHS) or LoF variants affecting *NSD2* and an antagonistic molecular consequence, it is necessary to differentiate this *NSD2*-related disorder and choose a disease name different from Rauch-Steindl syndrome.

## Data Availability

All data produced are available online at Zenodo (File S2,S3,S4: DOI: 10.5281/ZENODO.6206868).

https://doi.org/10.5281/ZENODO.6206868

## DATA AVAILABILITY

All data generated or analyzed during this study can be found either in the online version of this article at the publisher’s website or has been uploaded to Zenodo (File S2,S3,S4: DOI: 10.5281/ZENODO.6206868).

## ACKNOWLEDGEMENTS

We thank all involved families for participating in this study.

## AUTHORS’ CONTRIBUTIONS

B.P., M.B., T.Br., R.A.J. and V.S. conceived the initial study concept. B.P., M.B., M.W., T.Ba., R.A.J., T.Br. and V.S. analyzed genetic data. V.S., M.B. and T.Br. coordinated collection of clinical and genetic data. M.B., T.Br. and B.P. reviewed literature data and standardized the clinical HPO terms. M.B., S.P., M.R., N.D.D., J.G.A., E.G., M.W., T.Ba., J.H., T.M., R.A.J., T.Br. and V.S. provided clinical and genetic data and performed clinical assessments. B.P. performed structural protein analysis. B.P. analyzed all data and created all main figures. B.P., T.Br. and M.B. created the Supplementary files. B.P., M.B., V.S. and T.Br. wrote and edited the manuscript. All authors reviewed, commented and agreed on the final draft manuscript.

## SUPPLEMENTARY

**File S1** | Supplementary notes with supplementary figures and tables.

