## Supplementary notes for "A novel syndrome caused by the constitutional gain-of-function variant p.Glu1099Lys in *NSD2*"

### SUPPLEMENTARY METHODS

DNA Extraction from blood, buccal swabs and fingernails

DNA from EDTA blood was extracted using a chemagic™ 360 instrument (PerkinElmer) according to the manufacturer's recommendations. DNA from buccal swabs was extracted using DNA from buccal swabs (ORACollect DNA OCR-100, DNA Genotek) was extracted using the prepIT L2P reagent kit (DNA Genotek).

Sanger Sequencing

Polymerase chain reaction was performed according to standard procedures. DNA fragments were Sanger-sequenced using a SeqStudio Genetic Analyzer (ThermoFisher Scientific) according to the manufacturer's recommendations.

Exome Sequencing (ES)

Genomic DNA was extracted using standard methods from peripheral blood samples of Ind_1 and his parents. Trio exome sequencing and analysis was performed as described previously.[1,2] In brief, enrichment for exome sequencing of Ind_1 and both his parents was done using the BGI Exome capture 59M kit (BGI, Shenzhen, China) and sequenced with 100bp paired end reads on a BGISEQ-500 system (BGI, Shenzhen, China). The resulting sequencing data was processed using the cloud based "varfeed" pipeline (Limbus Medical Technologies GmbH, Rostock, Germany). The "varvis" webtool (Limbus Medical Technologies GmbH, Rostock, Germany) was used to filter the resulting variant files.

ES in Ind_2 was performed using genomic DNA which was extracted from leukocytes or whole blood using a chemagic 360 Instrument (PerkinElmer, US). SureSelect Human All Exon 60Mb V6 Kit (Agilent) was used for exome enrichment. Libraries were sequenced on an Illumina NovaSeq6000 system (Illumina, San Diego, California, USA) and reads were aligned to the UCSC human reference assembly (hg19) with BWA v.0.7.5a. On average more than 98% of targeted regions were covered at least 20x. Single-nucleotide variants (SNVs) and small insertions and deletions were detected using both SAMtools v.0.1.19 and GATK 4.1. Copy number variations (CNVs) were detected using ExomeDepth and Pindel. Variant prioritization was performed based on an autosomal recessive (minor allele frequency (MAF) <0.1%) and autosomal dominant (MAF <0.01%) inheritance.

Trio ES detected a heterozygous *de novo* missense variant in *NSD2* (NM_001042424.3:c.3295G>A, p.Glu1099Lys; chr4[hg19]:g.1962801G>A) in both individuals. No further candidate variants were prioritized.

Clinical data collection and comparison

The clinical terms in our Excel (Microsoft, Redmond, Washington, United States) based questionnaire were based on the phenotypes observed in Ind_1 and Ind_2 and a review of previous descriptions of individuals with RAUST [3] and standardized using the Human Phenotype Ontology (HPO) [4]. Clinical descriptions of patients with single nucleotide variants or indels in *NSD2* associated with RAUST were extracted from the reports or supplements of the primary literature source, individually reviewed and updated based on the published textual clinical descriptions or images. Fisher's exact test as implemented in R was used to compute p-values between GoF and likely LoF groups and p-values were corrected for multiple testing using false discovery rate (FDR) adjustment in R. Physical growth measurements were compared using World Health Organization (WHO) child growth standards using the pedz calculator (https://www.pedz.de/).

Variant spectrum and 3D structure analysis

The *NSD2* germline variants were harmonized to a common reference with VariantValidator [5] (NM_001042424.2 transcript, hg19 reference).
The distribution of NDD associated *NDS2* variants in the linear protein representation was compared to variants reported in the COSMIC cancer [6] database (file “V95_38_MUTANT.csv” downloaded on 2021-01-30) and in the public gnomAD [7] population database. Annotation, plotting and analysis of and protein regions constrained for missense variation was performed as described previously [8,9] with updated versions of all annotations.
For analysis of the missense variant distribution in the tertiary protein structure, we used the published Protein data bank (PDB) format structure 7CRO [10] of the SET and AWS domain of NSD2 together with the AlphaFold [11] homology model of human NSD2 downloaded from the EMBL website (https://alphafold.ebi.ac.uk/) with a pipeline based on the software PyMOL version 2.5.0 (Open-Source) as described before [8].

Analysis of omics data from cancer cell lines

We queried the Cancer Cell Line Encyclopedia (CCLE) [12,13] data using the “depmap” [14] and “ExperimentHub” packages in R version 4.1.0 from within RStudio version 1.4.1717. We used the metadata dataset "EH7294" together with the mutation calls ("EH7293") and copy number calls ("EH7291") to identify cell lines with NSD2 GoF/LoF or copy-number variants (CNVs) affecting the *NSD2* region. Cell lines with the known NSD2 GoF variants E1099K and T1150A were filtered using these annotation terms and classified into the GoF category while all nonsense or frameshifting variants were classified into the LoF category. Lines with a variant in *NSD2* not falling into these categories were classified as “other” and subsequently excluded from downstream analyses. The copy number data set was divided into five classes based on the log2 values ("duplication": log2 >= log2(3/2+1) ~ 1.32; "duplication purity 50%": log2 >= log2(5/4+1) ~ 1.17; "deletion purity 50%": log2 >= log2(3/4+1) ~ 0.81; "deletion": log2 >= log2(1/2+1) ~ 0.59; “neutral”). Cell lines with possible copy number changes at lower purity estimates were excluded from analysis.

To investigate chromatin modifications, we downloaded the “CCLE_GlobalChromatinProfiling_20181130” dataset directly from the depmap portal. The Z-score values of the chromatin marks at K27 and K36 were visualized as boxplots and group wise p-values were calculated using the “Wilcox rank sum test” as implemented in R.

To investigate differential methylation and differential expression, we next uploaded the lists of nine cell lines classified as GoF and 1192 lines classified as having no *NSD2* mutation and being copy number neutral (Supplementary File S02) to the depmap portal in the “Data Explorer” tool using the “Custom Analyses” tab.

We used the “Two class comparison” option to calculate the difference between two groups (GoF vs. neutral, excluding all cell lines with CNVs or any small *NSD2* variants) regarding both the methylation (“Methylation (1kb upsteam TSS)”) and the expression (“Expression 21Q4 Public”) dataset. Differentially methylated/ expressed gene lists were downloaded from depmap. Effect sizes, grouped by strength were plotted and -log10 scaled p-values were visualized as scatter plots using ggplot2 in R.

We filtered the list of differentially expressed genes to select 887 genes with a very high effect size (> 1.2 or < -1.2). This list was then uploaded to the humanbase [15] web tool (https://hb.flatironinstitute.org/) to predict functionally enriched gene modules with the “global” network setting.

### SUPPLEMENTARY RESULTS

Case of Individual 1

The boy Ind_1 (Fig. 1A) was the first child of healthy, non-consanguineous parents. The family history was uneventful for genetic disorders, intellectual disability, or seizures. Pregnancy was complicated by polyhydramnion and a prominent nuchal fold in first trimester ultrasound (US) screening. A horseshoe kidney was suspected in follow-up US examinations. Subsequent amniocentesis followed by conventional karyotyping and panel sequencing for Noonan syndrome were unremarkable. Due to premature rupture of the amniotic sac, pathological cardiotocography (CTG) and green discolored amniotic fluid, vacuum delivery was performed at 37+5 weeks gestational age. At birth his weight was 2,880 g (-0.85 SD), his length 48.0 cm (-1.22 SD), his occipital frontal circumference (OFC) 34.0 cm (-0.54 SD). Apgar scores were 8/9/9 and the umbilical cord pH was 7.09. Due to dyspnoea and decreased oxygen saturation he required intensive care with continuous positive airway pressure (CPAP) ventilation. His respiratory situation improved only slowly with continuous need for respiratory support. He required long-term total parenteral nutrition followed by duodenal tube feeding. Because of early cholestasis with conjugated hyperbilirubinemia, liver biopsy was performed, and histologic analysis of the liver tissue showed hepatocellular and canalicular cholestasis but normal intrahepatic bile ducts with no evidence of storage disease, viral inclusions, granulomata or steatosis. On his 21st day of life he developed sepsis and had to be intubated again for mechanical ventilation. Overall, he required prolonged intensive care.

Postnatally multiple organ anomalies were identified including persistent ductus arteriosus with aneurysm, splenomegaly with splenic cyst, cryptorchidism, bilateral dysplastic and enlarged kidneys with nephrocalcinosis, duplex kidney on the right side, bilateral vesicoureteral reflux and hypertrophic pyloric stenosis which required pyloromyotomy. He had preretinal hemorrhage of the left eye and astigmatism/ myopia were corrected using glasses. Hearing was normal. He had no seizures and electroencephalogram (EEG) was normal. Laboratory tests for etiological clarification of the suspected syndromic disease including metabolic testing were unremarkable.

He showed delayed developmental milestones. Formal IQ testing using the Snijders-Oomen non-verbale test (SON-R 2½-7) confirmed mild to moderate intellectual disability with an overall IQ of 50.

When the boy was last reviewed his height was 128.0 cm (-0.62 SD), his weight was 33.4 kg (1.01 SD; BMI 20,4), and his head circumference was 57.6 cm (3.47 SD). He presented with mild to moderate intellectual disability, could speak approximately five words and mainly communicated via gestures. Medical interventions regarding the malformations of the internal organs were not necessary. In particular, the laboratory parameters of renal and hepatic function were within the normal range. He had coarse facial features with square facial appearance and bitemporal narrowing, rather sparse and thin lateral eyebrows, low anterior hairline, multiple eyelid creases, periorbital fullness, narrow palpebral fissures, hypertelorism and convergent strabismus, a wide nasal bridge and a bulbous tip of nose, a long philtrum, a wide mouth, a thin upper lip with exaggerated cupid's bow, retrognathia and a short neck (Fig. 1A-D). His chest was broad with widely spaced nipples and the posterior thorax was broad with widely spaced scapulae and a flat vertebral column. The umbilicus was prominent after surgery for umbilical hernia. He showed a fixed elbow flexion and limited supination of the forearms. The hands had relatively broad palms, with brachydactyly and small nails and bridged palmar crease on the right side and single palmar crease on the left side (Fig. 1E-H). Similarly, the feet were short and broad, with bilateral pes valgus and hypoplastic nails (Fig. 1I-J). He had genu valgum and a broad-based and insecure gait (Fig. 1K). Examination of the external genitalia showed a micropenis.

Case of Individual 2

The boy Ind_2 (Fig. 1B) was the fourth child of non-consanguineous parents. Their family history was unremarkable for developmental disorders. Prenatal US revealed hepatomegaly, large echo-rich kidneys, right ventricular hypertrophy, unilateral clubfoot, macroglossia, single umbilical cord artery and polyhydramnios. Amniocentesis followed by prenatal karyotyping showed an unremarkable male karyotype (46,XY). High-throughput sequencing based panel testing for macrosomia and testing for Beckwith-Wiedemann Syndrome were negative. He was born preterm (33+4 gestational weeks, birth weight 2,740 g (1.9 SD), length 45.5 cm (0.47 SD), OFC 33.0 cm (1.13 SD)) via cesarean section due to missing fetal movements and pathological CTG. Apgar scores were 4/7/9, umbilical cord pH was 6.89. Acidosis resolved during the first 24 hours after birth. Respiratory insufficiency required intubation on the first day of life. Intracerebral hemorrhage of the right nucleus caudatus, grade 2 intraventricular hemorrhage, small cortical hemorrhages and signs of hypoxia of the occipital cortex were identified by MRI on day 9. Multiple congenital malformations were confirmed after birth including a congenital heart defect (hypoplastic aortic arch, ventricular septal defect), hepatomegaly, nephromegaly with small renal cysts, unilateral clubfoot (left foot), inguinal hernia (right side) and cryptorchidism. X-ray of the pelvis showed enlarged proximal femur metaphysis and prominent incisura ischiadica indicating skeletal dysplasia. A ventricular septal defect leading to increased right ventricular load was corrected via patch at the age of 5 weeks. Surgical repair of the hypoplastic aortic arch was performed. The club foot was treated by casting. Laboratory investigations revealed mild hyperammonemia (up to 150 µmol/l). Liver MRI identified a patent ductus venosus, which was treated by endovascular coiling. Repetitive episodes of respiratory distress treated by CPAP and oxygen therapy required hospitalization. Recurrent pyelonephritis with fever due to grade 2 vesicoureteral reflux were diagnosed and antibiotic prophylaxis was given. Nephrocalcinosis was identified by ultrasound but resolved spontaneously. Reduced counts of CD4+ T-cells and B-cells as well as increased numbers of NK-cells were identified; immunoglobulin levels (IgG and IgM) were borderline reduced. Severe recurrent infections were not present. Early on, deficits in head control and lifting the head and upper body while in prone prop position as well as muscular hypotonia were observed. Additionally, feeding difficulties and failure to thrive were prominent features. After the gastric tube was removed, he was able to eat soft foods and the BMI gradually normalized.

At the last investigation the individual had short stature and microcephaly (measurements: weight 12.0 kg (-1.46 SD; BMI 17,4), height 83.0 cm (-3.43 SD), OFC 46,5 cm (-3.32 SD)). Growth hormone stimulation test was abnormal. Hearing was noted to be impaired (failed BERA), most likely due to Eustachian tube dysfunction. Ophthalmologic assessment revealed myopia and astigmatism but was otherwise normal. Additionally, the individual presented with an eczema of unknown etiology of upper arms and legs as well as the face which did not resolve to treatment with steroids. Neurologic assessment revealed global developmental delay and a mild muscular hypotonia, but no signs of a movement disorder. He showed no clinical signs of seizures and EEG was normal. Motor development was markedly delayed. Cognitive and speech development was also delayed; the subject babbled repetitive syllables but was not able to speak any words. Evaluation for dysmorphic facial features identified multiple distinct features including thick hair, synophrys, long palpebral fissures with infraorbital crease, coarse facial features, depressed nasal bridge, anteverted nares, long philtrum, thin upper lip, microdontia, macroglossia, deep-set, and posteriorly rotated ears. Additionally, coarse and large hands with short fingers and a single transverse palmar crease as well as hypoplastic nails were present, while the feet were short and broad with hypoplastic nails. He had unilateral maldescensus testis.

**SUPPLEMENTARY FIGURES**

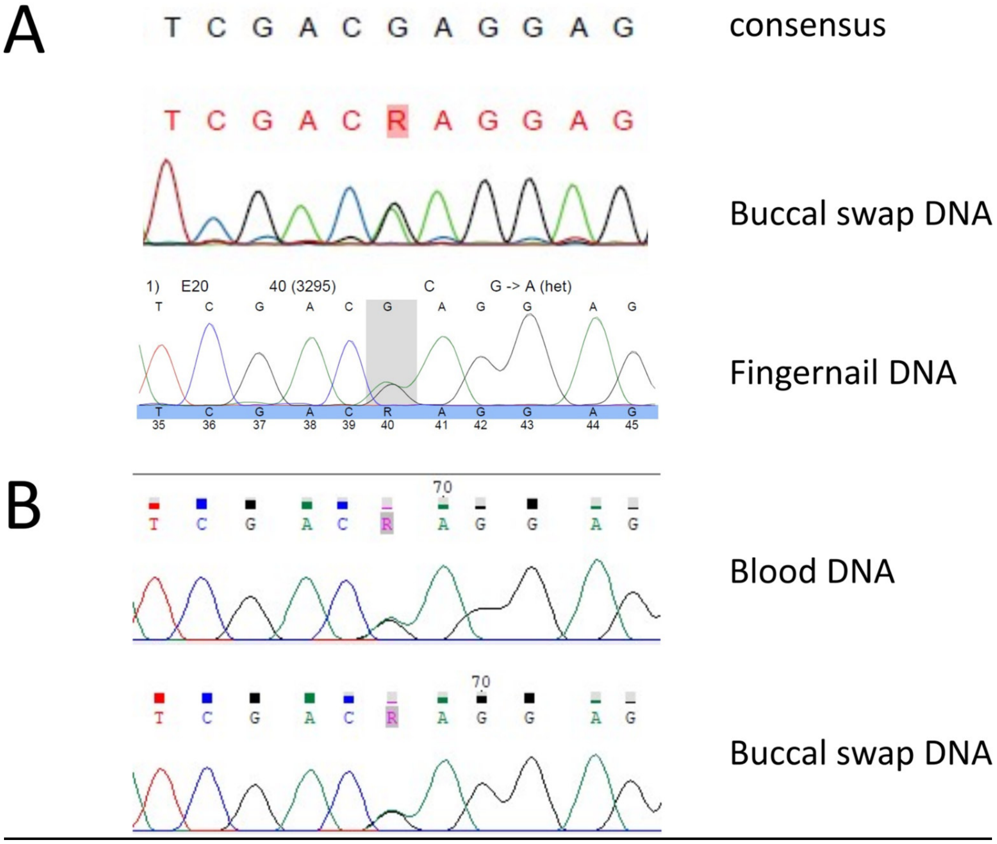


<u>**Figure S01** | Sanger sequencing of the missense variant c.3295G>A, p.Glu1099Lys in *NSD2* DNA from DNA from additional tissues.</u>

The missense variant in *NSD2* found in exome sequencing was confirmed in peripheral blood, buccal swab and fingernails in individual 1 **(A)** and in peripheral blood and buccal swab in individual 2 **(B)**, thus indicating germline origin.

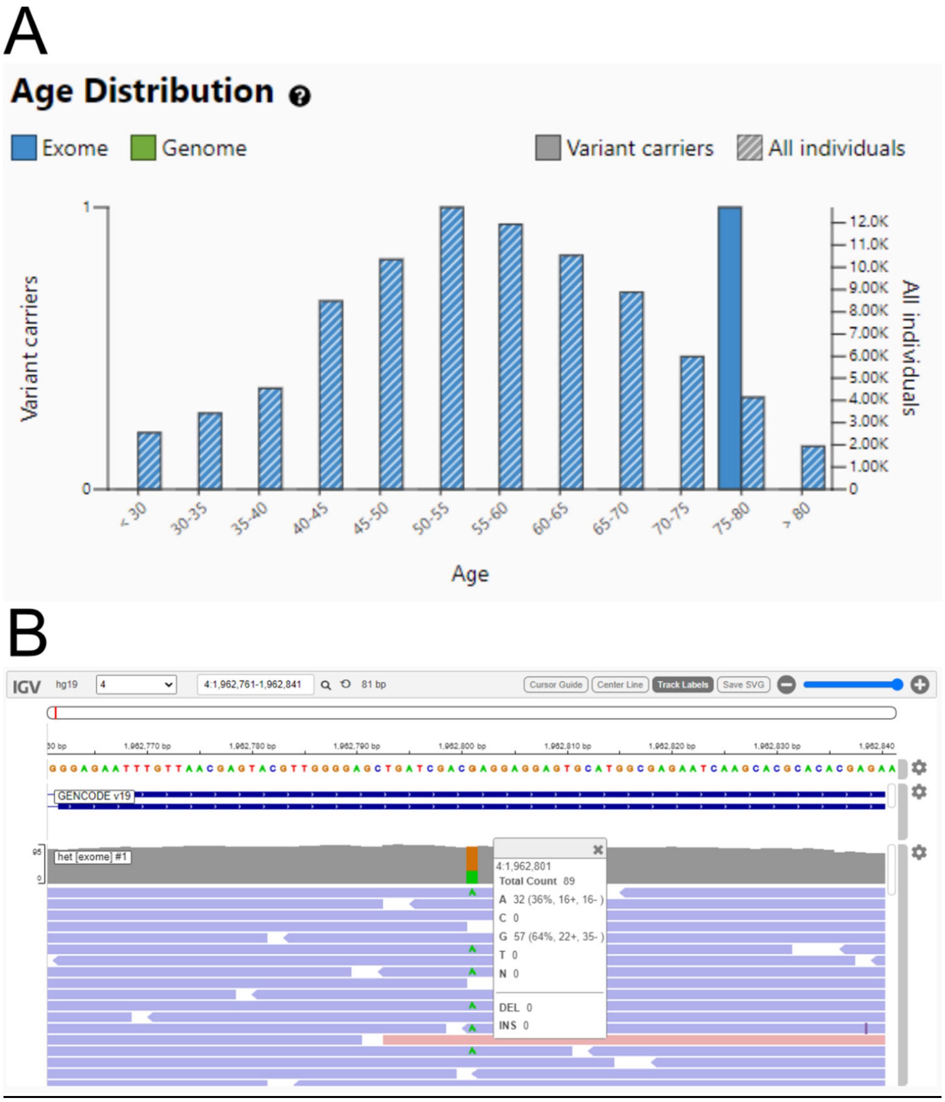


**Figure S02** | Somatic NSD2 variant c.3295G>A, p.Glu1099Lys in an elderly person

**(A)** Screenshot of the "Age Distribution" from gnomAD browser (https://gnomad.broadinstitute.org/variant/4-1962801-G-A?dataset=gnomad_r2_1) taken on 2021-01-30 showing that one of 4,136 individuals in the 75-80 years age range carries the c.3295G>A, p.Glu1099Lys *NSD2* variant. **(B)** Screenshot of the interactive IGV.js visualization of the read data of this individual indicates somatic status of the variant because of the skewed variant allele fraction with 32/96 (36%). Somatic driver mutations deriving from a unique hematopoietic cell lineage are regularly encountered in genomic databases of healthy populations such as gnomAD and pose a pitfall when misinterpreted as germline variant.[16] Diagnostic labs should be aware of this known pitfall and either use whitelists of somatic variants or allow a low allele count in *de novo* filters. [17]

### SUPPLEMENTARY DATA FILES

These data files used for analyses and tables/ figures are available for download from Zenodo:

**File S2 |** Clinical information of individuals described here and reviewed individuals together with all variant data.[18]

**File S3 |** Cell line analyses data.[18]

**File S4 |** humanbase functional modules data.[18]
